# Heterozygous truncating variants in *BICC1* are a novel cause of autosomal-dominant tubulointerstitial kidney disease

**DOI:** 10.64898/2026.08.20.26360556

**Authors:** Naomi Eylath, Kendrah O. Kidd, Piper Alyea-Herman, Julia Meyersiek, Dan A. Colombo, Helmut G. Rennke, Ava J. Guleserian, Victoria W. Adams, Giada Bianchi, Arnaud Maillard, Stanislas Faguer, Claudia Izzi, Carsten Bergmann, Stewart Lecker, Megan E Astley, Abbigail Taylor, Lauren Martin, Sydney Means, Antonio Sanchez, Nelson Weller, Kateřina Hodaňová, Tereza Kmochová, Viktor Stránecký, Hana Hartmannová, Klára Svojšová, Jakub Sikora, Lea Pavlovičová, Hana Yang, Peter C. Harris, Stanislav Kmoch, Anthony J. Bleyer, Martina Živná, Peter G. Czarnecki

## Abstract

**Introduction:** Autosomal-dominant tubulointerstitial kidney disease (ADTKD) is characterized by chronic kidney disease (CKD) with an average age of end-stage renal disease (ESRD) of approximately 45 years, bland urinary sediment, the absence of proteinuria and autosomal dominant inheritance. While several causative genes have been found, there remain families in whom no molecular diagnosis has been identified (ADTKD-NMD).

**Methods:** We identified *BICC1* truncating variants in several families with ADTKD-NMD in the Wake Forest Rare Inherited Kidney Disease Registry and then screened families in our database and other referred families for *BICC1* truncating variants. We performed segregation analysis and characterized affected individuals for clinical and histopathologic phenotypes. We analyzed oligomer formation of BICC1 mutants with wild-type BICC1-, ANKS3- and ANKS6 proteins through co-immunoprecipitation and Western blotting, and we tested for posttranscriptional regulation of the BICC1 target mRNA, *Dand5*, in a Luciferase reporter assay.

**Results:** We found 6 heterozygous truncating mutations in *BICC1* segregating with the ADTKD phenotype in 8 independent pedigrees worldwide. Affected individuals developed kidney failure in the 6^th^ to 7^th^ decade of life that was characterized pathologically by tubular atrophy and interstitial fibrosis. The truncated gene products localized to cytoplasmic bodies and demonstrated various degrees of self-association or binding to the known interaction partners, ANKS3 and ANKS6. While the wild-type *BICC1* gene product acts as a posttranscriptional repressor of target mRNAs, all truncation variants exhibited increased expression of substrate mRNA.

**Conclusions:** Truncating variants in *BICC1* are a novel cause of ADTKD, segregating with the disease phenotype and upregulating *BICC1* target gene expression through a dominant-negative- or a gain-of-function mode of action.

**Translational Statement:** Novel disease gene discoveries have a high potential for translational impact. Autosomal-dominant tubulointerstitial kidney disease (ADTKD) is reported to occur in 1-2 individuals per 100,000 but is underrecognized and underdiagnosed. Although several genes have been associated with ADTKD, many cases remain genetically unresolved. Our discovery of *BICC1* as a novel gene in ADTKD will lead to improved disease recognition, prognostication and counseling. Through its role in gene regulation, BICC1 is a *bona fide* target for the molecular study of kidney fibrosis and atrophy, a final common pathway in CKD, as well as for the development of future therapies.

## Introduction

Autosomal-dominant tubulointerstitial kidney disease (ADTKD) is a monogenic disorder characterized by progressive chronic kidney disease, a bland urinary sediment and tubulointerstitial fibrosis. Affected individuals develop kidney failure between 30 and 80 years of age. Since ADTKD lacks distinctive clinico-pathologic features, its diagnosis relies on family history and genetic testing. Therefore, ADTKD is widely underrecognized and underdiagnosed, and the true disease burden is likely higher than the reported prevalence of 1-2/100,000^1^. Pathogenic variants in *UMOD*, *MUC1* and *REN* are well-characterized causes of ADTKD. ADTKD-like phenotypes may also be seen in carriers of heterozygous mutations in *HNF1B, JAG1, NOTCH2, DNAJB11* and other genes, where phenotypic overlap with cystic- and congenital anomalies of the kidney and urinary tract (CAKUT)-spectrum disorders is common^1-3^. In slowly progressive disease, the diagnosis of ADTKD is likely to be missed because patients present at an advanced age, when other, non-genetic causes of CKD are common, and when family histories are less comprehensively collected. In this study, we report the discovery of truncating variants in the Bicaudal-C1 gene, *BICC1,* as a novel cause of ADTKD. We identified this gene through enrichment of heterozygous pLoF variants in the *BICC1* gene, and their segregation with a progressive CKD phenotype in well-documented pedigrees affected by ADTKD-NMD. *BICC1* is a pleiotropic gene involved in kidney development and has previously been associated with recessive CAKUT in humans^4^, as well as with recessive cystic kidney disease in mice^5^. To our knowledge, this is the first report of dominant *BICC1*-related kidney disease, and we present various heterozygous truncation alleles from unrelated pedigrees around the world, all of which manifest with the same isolated ADTKD phenotype.

## Materials and Methods

### Chart review and clinical-/pathologic data

This study was approved by the institutional review boards of Wake Forest University Health Sciences (Winston-Salem, NC, United States; IRB #00000352) and Charles University (Prague, Czech Republic). We obtained demographic data, laboratory results and family histories. Blood- or saliva samples from study participants were processed for genomic DNA extraction and whole exome sequencing, as previously described^6^.

### Plasmids and molecular cloning

*Renilla reniformis* Luciferase cDNA was PCR amplified using the plasmid pIRIGF as a template, and inserted into pcDNA3.1(+) using NheI/BamHI, omitting the stop codon. All other pcDNA3.1(+)-derived expression vectors carry 3xFLAG-, 3xmyc- or StrepTags, positioned between the NheI and BamHI sites, as previously described^7^. Wild-type *BICC1* cDNA was amplified by PCR and subcloned into pcDNA3.1(+) vectors with BamHI/NotI, generating N-terminal *Renilla* Luciferase, 3xFLAG-, 3xmyc- or StrepTag fusion constructs, respectively. All truncation variants were introduced through PCR-induced mutagenesis. For the generation of lentiviral expression vectors, 3xFLAG-tagged cDNA constructs were subcloned into pCDH-UbC-MCS-EF1A-Puro, using NheI/NotI. *Rattus norvegicus Anks6* and *Mus musculus Anks3* cDNA constructs were subcloned into the respective pcDNA3.1 expression vectors through PCR amplification and BamHI/NotI cleavage. The firefly luciferase reporter vector was cloned by PCR amplification of *Photinus pyralis* Luciferase cDNA containing its stop codon, using pIRIGF plasmid as a template, and inserting into pcDNA3.1(+) using NheI/BamHI. Subsequently, a 144 bp fragment of *Xenopus tropicalis Dand5* 3’UTR was amplified by PCR and inserted downstream of the Luciferase sequence using BamHI and KpnI sites. Small-guide DNA oligonucleotides targeting *Mus musculus Bicc1* were annealed and ligated into the BbsI site of pSpCas9(BB)-2A-GFP. All DNA modifying enzymes were obtained from New England BioLabs. All oligonucleotides were synthesized by IDT, with sequences listed in the supplemental materials section. All expression vectors were verified through next-generation sequencing of the entire plasmid (Plasmid-EZ, Azenta).

### Cell culture, transfection and lentiviral transduction

HEK293T cells were cultured in DMEM; mIMCD-3 cells were cultured in DMEM/F-12; each supplemented with 10% FBS and Penicillin/Streptomycin. For transient transfection, HEK293T cells were plated in 10-cm petri dishes and transfected with 6 μg DNA and 18 μg PEI in 250 μl OptiMEM. For packaging of lentiviral particles, HEK293T cells were grown in 25 cm^2^ flasks and transiently transfected with a mix of 2 μg lentiviral vector, 1 μg psPax2, 1 μg pMD2.G, and 12 μg PEI. After 24 h, the supernatant containing lentiviral particles was collected, filtered through a 0.45 μm syringe filter, supplemented with 7 μg ml^-1^ polybrene, and applied to mIMCD-3 cells in 25 cm^2^ flasks. Media were changed to fresh DMEM/F-12/10% FBS, and selection with 1 μg ml^-1^ puromycin was begun 24 h after application of lentivirus.

### CRISPR/CasG-mediated knockout of *Bicc1*

mIMCD-3 cells were grown to 50% confluence in a 25 cm^2^ flask and transiently transfected with 4 μg pSpCas9(BB)-2A-GFP vector harboring the *Bicc1* sgDNA. Cells were trypsinized and spun down 48 h post transfection, resuspended in PBS with 2% FBS, labeled with DAPI 5 μg ml^-1^, and subjected to FACS sorting on a BD FACS Aria sorter. We sorted 1 cell per well in 96-well plates, gating for doublet exclusion, DAPI exclusion and GFP positivity. Monoclonal cell lines were genotyped through genomic DNA extraction (DNeasy Blood and tissue kit, QIAGEN), amplification of the targeted *Bicc1* Exon 1 through two rounds of nested PCR, and next-generation sequencing of the amplicon using the Amplicon-EZ platform (Azenta).

### Protein pulldown/immunoprecipitation

HEK293T cells were collected 24 h post transient transfection, and lyzed in 25 mM Tris-HCl pH 7.5, 150 mM NaCl, 1% Triton X-100. Lysates were clarified by centrifugation, protein concentrations were measured with a modified Bradford assay, and samples were diluted with lysis buffer to 500 μg ml^-1^. Proteins were captured with anti-myc-, or StrepTactin®XT-4Flow agarose beads, washed 4 times with 1 ml lysis buffer, and eluted with Laemmli buffer or 50 mM biotin in lysis buffer.

### Western blotting

Protein samples (whole cell lysates or eluates from immunoprecipitation) were supplemented with Laemmli buffer and separated on hand-casted Mini-PROTEAN SDS-PAGE gels (Bio Rad) in a Laemmli SDS/Tris-Glycine buffer system. Gels were blotted onto Nitrocellulose membrane, blocked in 2.5% bovine serum albumin in TBST and incubated with the respective antibodies. Signals were detected on X-ray films through ECL.

### Dual-luciferase assay

mIMCD-3 Δ*Bicc1* #B1B3 cells, or HEK293T cells were transfected with a mix of the respective *Renilla reniformis* Luciferase-tagged *BICC1* construct and *Photinus* luciferase-*Dand5*-3’UTR reporter plasmid. After 24 h, dual luciferase assay was conducted using the Promega Dual-glo® assay kit, following the manufacturer’s protocol. We used 1 s signal integration time for both, *Renilla* and *Photinus* luciferase activity measurements. BICC1 functional activity was expressed as a ratio of *Photinus* and *Renilla* luciferase relative light emission units.

### Immunofluorescence microscopy

Wild-type and *Bicc1* CRISPR/cas9 knockout mIMCD-3 cells were lentivirally transduced with the respective *BICC1* cDNAs and grown on sterile 18 mm coverslips in 12-well plates. Cells were fixed in 4% formaldehyde in PBS for 10 min, quenched in 300 mM ethanolamine pH 9.0 for 5 min, permeabilized with 0.4% Triton X-100 in PBS for 5 min, and blocked in 2.5% bovine serum albumin (BSA) in PBS with 0.05% Triton X-100 for 1 h. A carrier solution of 1% BSA in PBS with 0.05% Triton X-100 was used for all antibodies and subsequent washes. Antibodies were incubated for 1 h each, followed by three subsequent washes. Cells were incubated in DAPI at 5 μg ml^-1^, washed two times with MilliQ water and two more times with 100% ethanol. Coverslips were dried and mounted in Vectashield (Vector Labs). Images were acquired with a Zeiss LSM 880 upright confocal microscope system and analyzed with the ImageJ software package.

### Immunohistochemistry

Formaldehyde-fixed, paraffin-embedded (FFPE) kidney biopsy tissue was deparaffinized and rehydrated using standard protocols. Endogenous peroxidase activity was blocked by incubation in 1% sodium azide and 0.3% hydrogen peroxide for 10 minutes. Antigen retrieval was performed according to Okada K et al^8^. BICC1 protein was visualized through incubation with rabbit polyclonal anti-human BICC1 antibody and EnVision+ System-HRP (Dako). Sections were counterstained with hematoxylin.

### Statistical analysis

Standard statistical analyses were performed with SAS (SAS Institute). Estimated glomerular filtration rate (eGFR) was calculated using the CKD-EPI 2009 equation^9^. These values were compared to age- and sex-specific reference values^10^. Luciferase assay data were analyzed by Welch’s t-test (Prism 11 for MacOS, GraphPad).

## Results

### Identification of a heterozygous *BICC1* variant in a pedigree with clinical ADTKD-NMD

The index case (**Figure 1A**, III.2) was a male in his 50s with an eGFR of 49 ml/(min * 1.73 m^2^), presenting for the evaluation of chronic kidney disease (CKD) of uncertain etiology. Urinalysis revealed a bland urinary sediment, and the urine albumin-to-creatinine ratio was 110 mg/g. A kidney ultrasound revealed an 11 cm right kidney and a 12.1 left kidney without cysts. Family history (see **Figure 1A**) revealed that the index case’s parent (II.1) started hemodialysis in her 60s due to CKD of unknown cause. Their grandparent (I.1) died in their 70s from kidney failure without renal replacement therapy. One sibling (III.1) developed kidney failure of unknown cause in his 60s, and another sibling (III.5) suffered from CKD of unknown cause with an eGFR of 27 ml/(min * 1.73 m^2^) also in his 60s. He also had well-controlled type 2 diabetes mellitus, as well as coronary and cerebrovascular disease. His urinalysis revealed 1+ protein. Another sibling (III.4) had an eGFR of 61 ml/(min * 1.73 m^2^) in her 50s. In summary, the family pedigree (**Figure 1A**) i s suggestive of CKD of genetic origin with an autosomal dominant mode of inheritance. Natera Renas ight^©^ kidney disease gene panel testing was negative for variants in the known genes causing autosomal dominant tubulointerstitial kidney disease, but revealed a heterozygous truncating variant in *BICC1,* c.1977_1978del, p.Val660Glyfs*15 in two affected siblings (see **Table 1**). Separate genetic test ing for *MUC1* pathogenic variants was negative^11-13^. Further systematic genotyping by whole exome sequencing confirmed the same *BICC1* variant in all four affected siblings, while excluding other shared potentially pathogenic variants. The Wake Forest Team hypothesized that this and potentially other heterozygous variants in *BICC1* may represent a novel cause of autosomal-dominant tubulointerstitial kidney disease.

**Figure 1.**
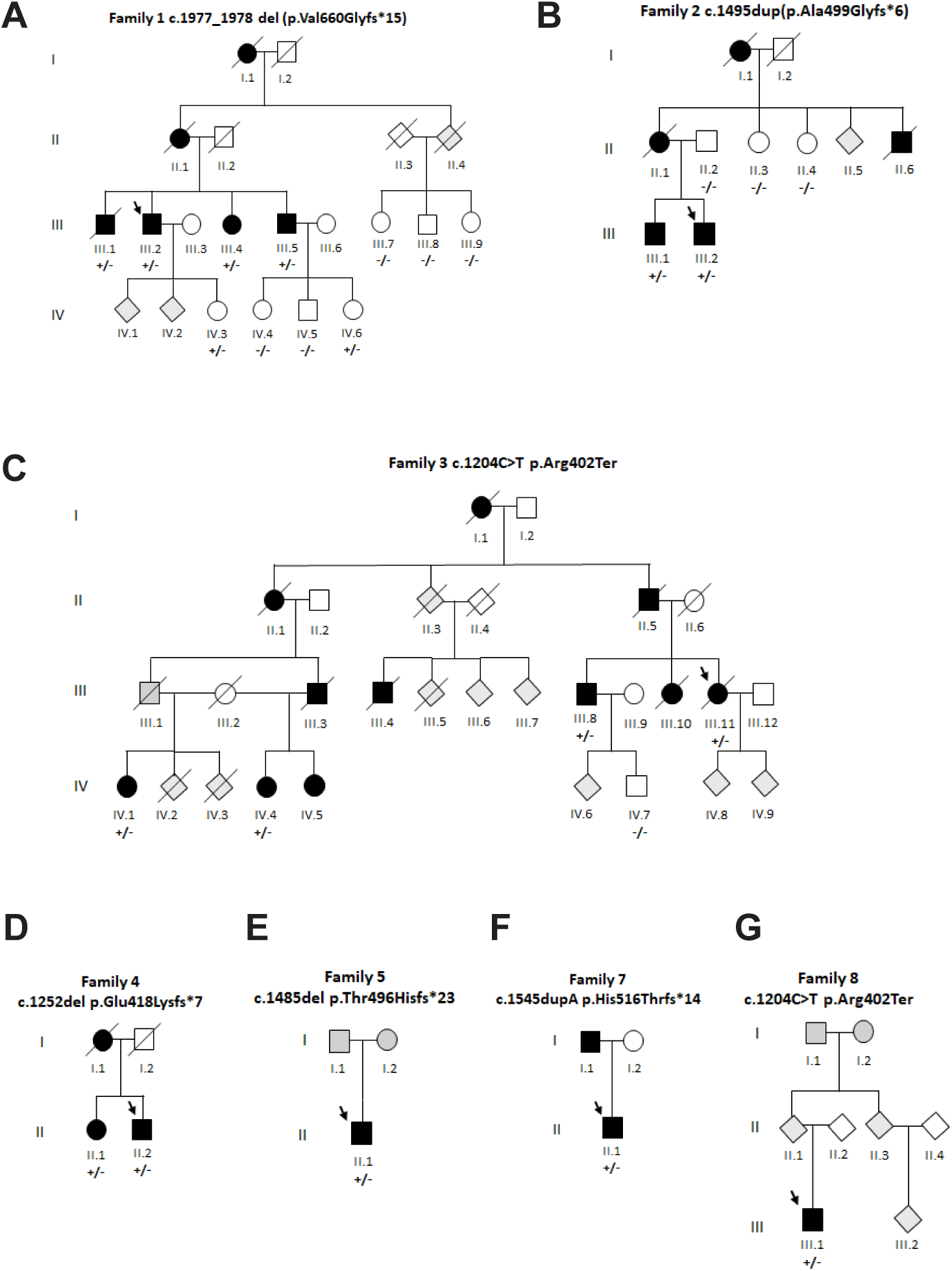
Pedigrees affected by ADTKD-*BICC1*. **A–** The index case leading to the discovery of *BICC1* as a novel ADTKD gene is individual III.2 in Family 1. Family members at risk in generation III were completely genotyped and demonstrate complete genotype-phenotype correlation of the *BICC1* c.1977_1978del variant with CKD/KF. Individuals IV.3 and IV.6 carry the pathogenic variant, but did not develop disease, presumably because of young age. **B–** Family 2 carries the *BICC1* c.1495dup variant and demonstrates complete genotype-phenotype correlation. **C–** Family 3 carries the *BICC1* c.1204C>T variant. Not all individuals at risk were available for genotyping. **D–F–** Families 4, 5 and 7 carry the *BICC1* variants c.1252del, c.1486del and c.1545dup and demonstrate genotype-phenotype correlation in all tested individuals. **G–** Family 8 is affected by the *BICC1*c.1204C>T variant and demonstrates genotype-phenotype correlation in all tested individuals. The missense variant is identical with the one in Family 3 and with a singleton (Family 6, not depicted here), but long-range PCR reveals different patterns of flanking single-nucleotide polymorphisms between the three kindreds, demonstrating allelic heterogeneity (Supplemental Figure S1).

**Table 1.**
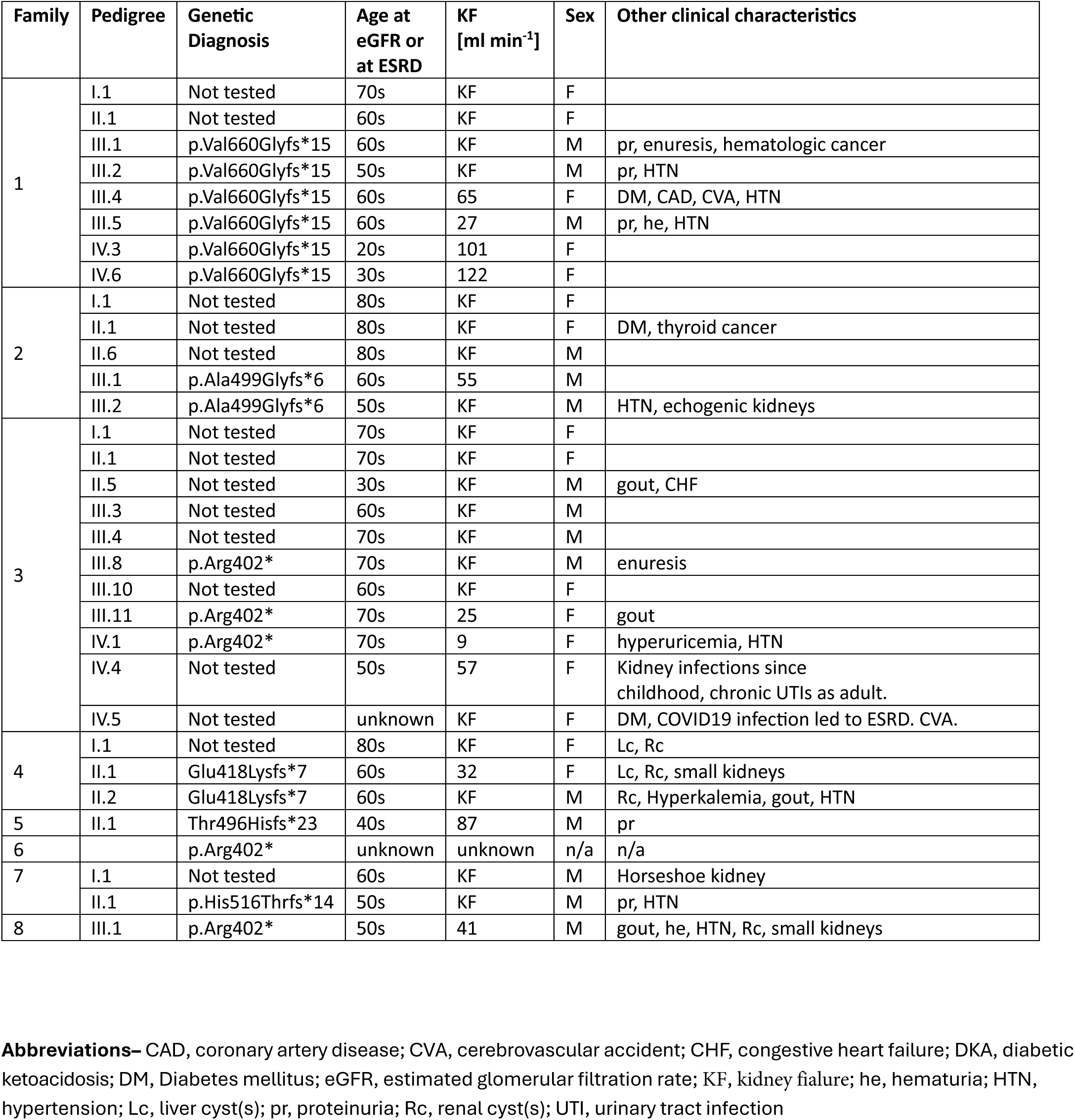
Clinical characteristics of *BICC1* pLOF variant carriers.

### Heterozygous truncating variants in *BICC1* segregate with ADTKD phenotypes in multiple pedigrees

Further evaluation of 211 cases from the Wake Forest Rare Inherited Kidney Disease Registry with a Natera Renasight® report on file revealed 85 cases with no identified genetic cause. An additional 62 cases underwent genetic testing on the Mayo Clinic cystic kidney disease and ciliopathy gene panel. We found the presence of heterozygous truncating *BICC1* variants in seven additional families, bringing the total to eight. The variant c.1495dup, p.Ala499Glyfs*6 in the index case of Family 2 (III.2, **Figure 1B**), was identified through Natera Renasight^©^ panel testing, and the heterozygous variant c.1204C>T, p.Arg402*, in the index case of Family 3 (III.11, **Figure 1C**), was identified through the Mayo Clinic panel^14^. A query in GeneMatcher^15^ returned nine submitting institutions reporting variants in *BICC1*. One team contacted us with three identified families in which heterozygous truncating *BICC1* variants segregated with an ADTKD phenotype (Family 4, c.1252del, p.Glu418Lysfs*7, **Figure 1D**; Family 5, c.1486del, p.Thr496Hisfs*23, **Figure 1E**; and Family 7, c.1545dup p.His516Thrfs*14, **Figure 1F**). Additional families 6 (complete pedigree not available) and 8 (**Figure 1G**) were found to carry the *BICC1,* c.1204C>T, p.Arg402* variant, through ClinVar submission^16^ (Accession: VCV003236256.2) and through personal communication with their treating physicians, respectively. Families 3, 6 and 8 carried the same 1204C>T, p.Arg402* variant. Importantly, long-read sequencing across the genomic locus revealed different flanking single nucleotide polymorphisms, indicating that the identical stop-gain variant arose independently in three different kindreds, and suggesting that the c.1204C>T site represents a recurrent mutational hotspot (**Supplemental figure S1**). **Tables 1 and 2** summarize demographic and clinical/ pathologic data of our study subjects (see also **Supplemental Table 1**). 25/30 individuals had abnormal kidney function (eGFR ≥2 s.d. from the mean eGFR based on age and gender^11^, compared with 1/6 family members without *BICC1* mutations (p=0.000874). From 65 patients at risk in 8 distinct pedigrees, 32 were considered to have ADTKD-*BICC1*, either based on a confirmatory genetic test (n=17), or, if genetic testing was not available, based on clinical suspicion in the presence of CKD/KF and plausible autosomal-dominant segregation (n=15). Of the 17 patients testing positive for a pathogenic variant, 15 had CKD or KF (age range 43-71), and the two remaining patients have likely not yet developed clinically significant disease (ages within 16-35). Among the 32 patients with proven or suspected ADTKD-*BICC1*, 5 had m ild proteinuria, 5 had hyperuricemia or gout, 4 had small kidney cysts, 2 had liver cysts, and 2 had CAKUT presentations (recurrent urinary tract infections beginning in childhood, horseshoe kidney). In 18 patients with KF and available clinical and demographic information, the median age at KF was 70 years (range, 39 to 83 years). **Figure 2** depicts eGFR as a function of age for the entire study population.

**Figure 2.**
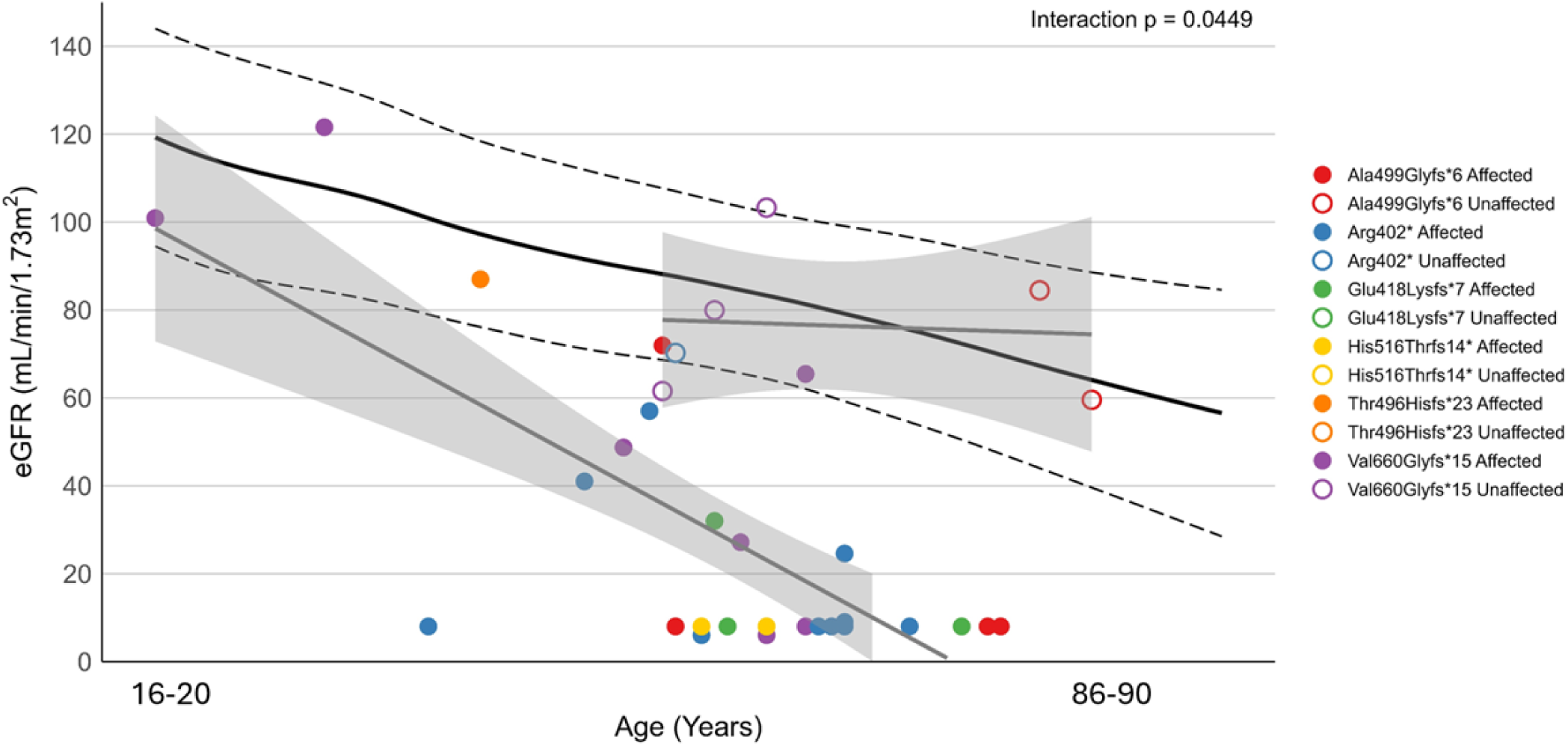
Progressive kidney function decline in *BICC1* wild type and truncating variant carriers. Estimated glomerular filtration rate, calculated using the CKD-EPI 2009 equation^9^ for individuals with *BICC1* pathogenic variants (darkened circles) and unaffected family members (open circles). 1/6 unaffected individuals had mild CKD, vs. 25/30 genetically affected individuals having abnormal kidney function (p=0.000874). The 95% confidence intervals are from age- and sex-specific reference values from a cohort consisting of 1.5 million healthy Europeans from the European Chronic Kidney Disease Burden Consortium^10^.

**Table 2.** Genotypes, phemotypes and classification of identified *BICC1* predicted loss of function (pLOF) variants.

| Family | Ancestry/<br>Nationality | Inheritance | Onset | Renal Phenotype | Nucleotide<br>change | Amino Acid<br>Change | Zygosity | Franklin<br>by<br>Genoox<sup>16</sup> | ACMG<sup>17</sup> | Allelic<br>frequency<br>(gnomAD<br>v4.1.1<sup>18</sup>) | Variant ID<br>(ClinVar<sup>15</sup>) |
| --- | --- | --- | --- | --- | --- | --- | --- | --- | --- | --- | --- |
| **1** | Russian | AD Maternal | Adult | CKD, ESRD, pr, he, DM,<br>HTN, enuresis, CAKUT | c.1977_1978del | p.Val660Glyfs*15<br>(frame shift) | het | LP | PVS1,<br>PM2 | n/a | 3596161 |
| **2** | Italian | AD Maternal | Adult | CKD, ESRD, DM | c.1495dup | p.Ala499Glyfs*6<br>(frame shift) | het | LP | PVS1,<br>PM2 | n/a | n/a |
| **3, 6, 8** | Irish (3)<br>n/a (6)<br>Chinese (8) | AD<br>Paternal (3)<br>n/a (6)<br>Maternal (8) | Adult | CKD, ESRD, gout, DM,<br>CAKUT, enuresis (3)<br>n/a (6)<br>Gout, HTN, CKD, DM (8) | c.1204C>T | p.Arg402*<br>(stop gain) | het | LP | PS4,<br>PVS1,<br>PM2,<br>PP5 | 2.48e-6 | 3236256 |
| **4** | French | AD Maternal | Adult | CKD, hyperkalemia, Rc,<br>Lc | c.1252del | p.Glu418Lysfs*7<br>(frame shift) | het | LP | PVS1,<br>PM2 | n/a | n/a |
| **5** | French | Singleton | Adult | CKD, pr | c.1486del | p.Thr496Hisfs*23<br>(frame shift) | het | LP | PVS1,<br>PM2 | n/a | n/a |
| **7** | French | AD Paternal | Adult | HTN, ESRD, CAKUT | c.1545dupA | p.His516Thrfs*14<br>(frame shift) | het | LP | PVS1,<br>PM2 | n/a | n/a |

**Abbreviations:** AD, autosomal dominant; CAKUT, congenital anomaly of kidneys and urinary tract; CKD, Chronic Kidney Disease; DM, diabetes mellitus; ESRD, End-stage Renal Disease; he, hematuria; het, heterozygous (het); HTN, hypertension; Lc, Liver cyst(s); LP, Likely Pathogenic; PM2, Population data extremely low allelic frequency; PP5, Reputable source of pathogenicity; PS4, Population Data higher prevalence of a rare variant in unrelated patients with the same phenotype than in healthy controls; pr, proteinuria; PVS1, Functional data null variant; Rc, Renal cyst(s)

### Histopathology

We reviewed the histopathologic findings in three patients who underwent kidney biopsy as part of their diagnostic workup, and we found tubular atrophy, interstitial fibrosis and glomerular obsolescence beyond the expected level for the given age. Interstitial fibrosis was accompanied by chronic inflammation with infiltration by mononuclear cells (**Figure 3**). Some tubules revealed microcystic changes, with cysts containing PAS-positive secretion product. Glomerular and tubular hypertrophy represented compensatory adaptations. Similarly, focal segmental or global glomerulosclerosis were regarded as secondary lesions indicating nephron dropout, while no primary glomerular injury was present. As in ADTKD-*UMOD* or ADTKD-*MUC1*, the histopathology lacked distinctive pathognomonic features, and the same histologic changes could be observed in the context of CKD of various other etiologies.

**Figure 3.**
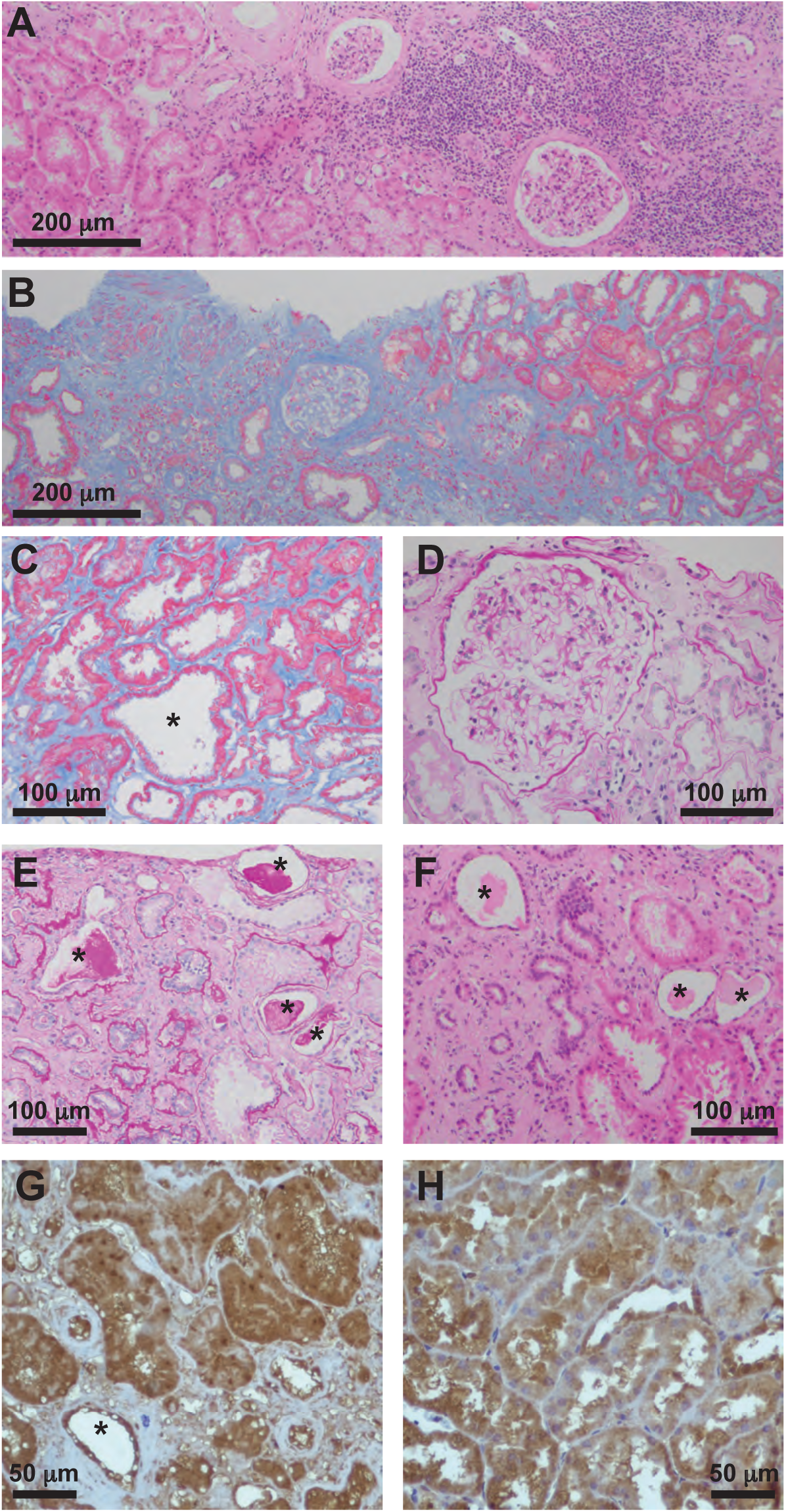
Histopathologic findings of ADTKD-BICC1. **A–** Hematoxylin-Eosin-(H/E)-stained section of a kidney biopsy specimen from a patient with ADTKD-BICC1. Widespread tubular atrophy with areas of interstitial inflammation and infiltration by mononuclear cells; **B–** Masson’s Trichrome stain reveals the extent of underlying interstitial fibrosis; **C–** Higher magnification of the Trichrome-stained section shows atrophic tubules, sometimes with cystic dilatation, and extracellular matrix expansion with peritubular fibrous deposition; **D–** Periodic-acid-Schiff-(PAS)-stained section depicts glomerular hypertrophy with segmental sclerosis, as an expression of hyperfiltration and nephron dropout; **E–F–** PAS- and H/E-stained detail of areas of tubular atrophy with cystic dilatation and PAS-positive secretion product. **G–H–** Immunoperoxidase-stained sections of biopsy specimens of a patient with ADTKD-BICC1 (G) and a healthy control (H), using anti-BICC1 antibody. BICC1 is predominantly expressed in tubular epithelial cells.

### Structure-function analysis and subcellular localization

**Figure 4** illustrates the domain architecture of the Bicaudal-C1 protein, comprising an N-terminal region (amino acids 45-353) with three K-homology domains (KH1-3), a C-terminal region containing a sterile alpha motif- (SAM-)domain (amino acids 871-936), and an intervening linker region devoid of any predicted structural features. While the KH-domains function through binding to specific RNA substrates, the SAM-domain has been implicated in the structural organization of BICC1 protein into multimers, and for its interactions with other SAM-domain containing proteins, like ANKS3 and ANKS6^20^. Determinants within the unstructured linker region mediate posttranscriptional repression of the bound RNA substrates^21^. All identified truncating variants are located within the linker region and are predicted to result in the loss of the SAM-domain (**Figure 4**). We next investigated whether truncated BICC1 protein fragments could be stably expressed in renal tubular epithelial cells. To this end, we transduced mouse IMCD-3 cells with lentiviral particles carrying either wild-type or truncation variants of *BICC1* cDNA. BICC1 protein was previously described to fulfill its biological function within cytoplasmic P-bodies/ RNA processing granules^22,23^. Therefore, we sought to determine the subcellular localization of the truncated BICC1 protein isoforms in comparison to the wild-type protein and performed immunofluorescence analysis of the lentivirally transduced IMCD-3 cells. As shown in **Figure 5A–B**, wild-type BICC1 protein distributed mostly homogenously throughout the cytosol of IMCD-3 cells, with occasional granules. In contrast, most truncated variants demonstrated a speckled pattern, reminiscent of cytoplasmic granules reported previously^22,24^. Remarkably, all truncation variants demonstrated a strong granular localization when expressed in mIMCD-3 cells in which the endogenous *Bicc1* locus was knocked out by CRISPR/Cas9, and in which the transduced *BICC1* variant represented the only isoform present. However, when expressed in native mIMCD-3 cells that contain endogenous full-length Bicc1 protein, all truncated isoforms localized less strongly to granules and distributed more evenly throughout the cytoplasm.

**Figure 4.**
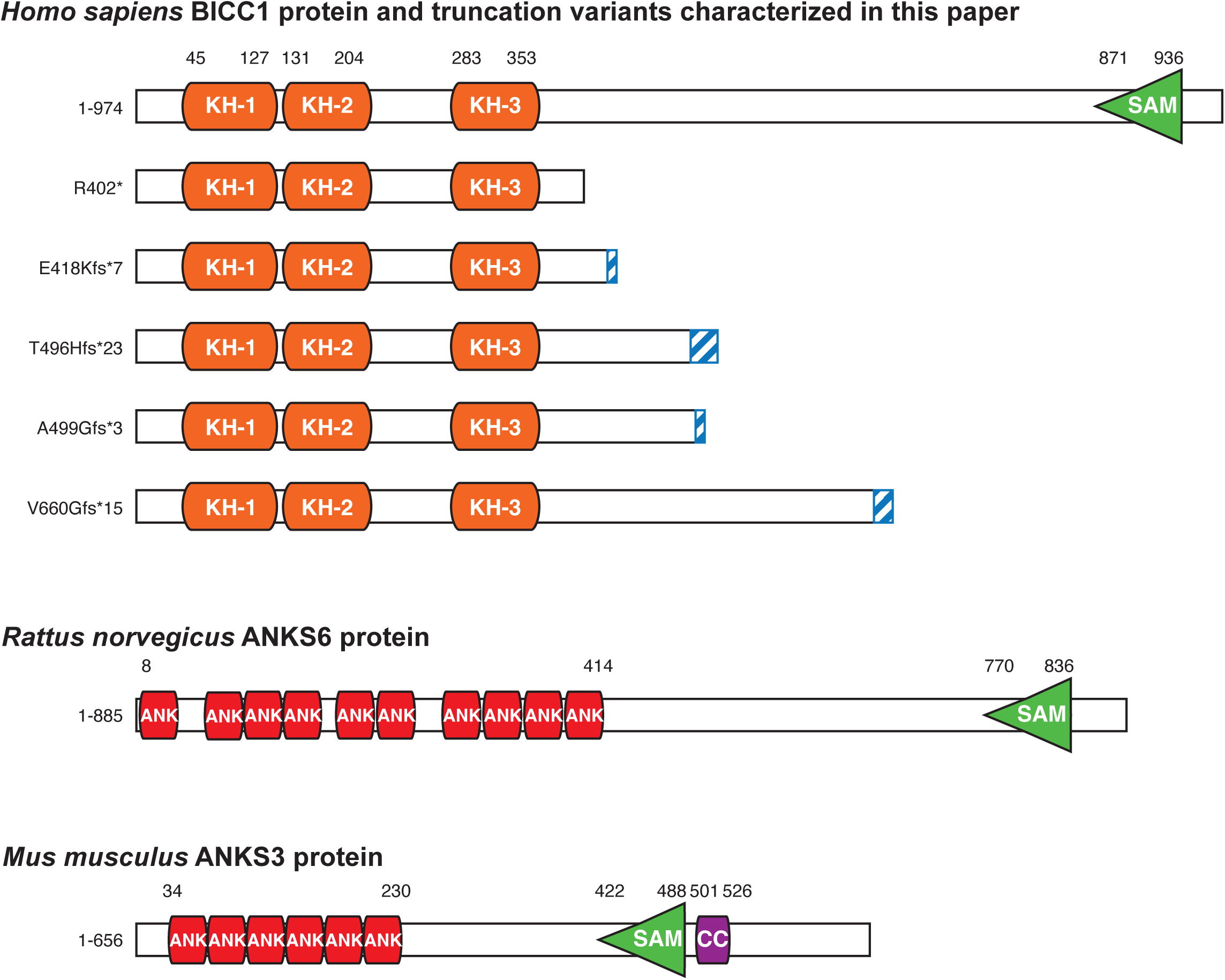
Protein domain architecture of wildtype and truncated *BICC1* gene products, and the related SAM-domain proteins ANKS3 and ANKS6. Full-length BICC1 protein contains an array of three K-homology domains in its N-terminal region, and a C-terminal sterile-alpha-motif-(SAM)-domain. It is unknown if the linker region (amino acids 353-871) assumes any organized secondary- or tertiary structure fold. All variants except for the stop/gain-variant R402X result in the addition of a random C-terminal extension as a result of a frame shifting event. The related SAM-domain proteins ANKS3 and ANKS6 are interaction partners that are being recruited to BICC1 oligomers, and that exhibit a similar modular composition with C-terminal SAM-domains.

**Figure 5.**
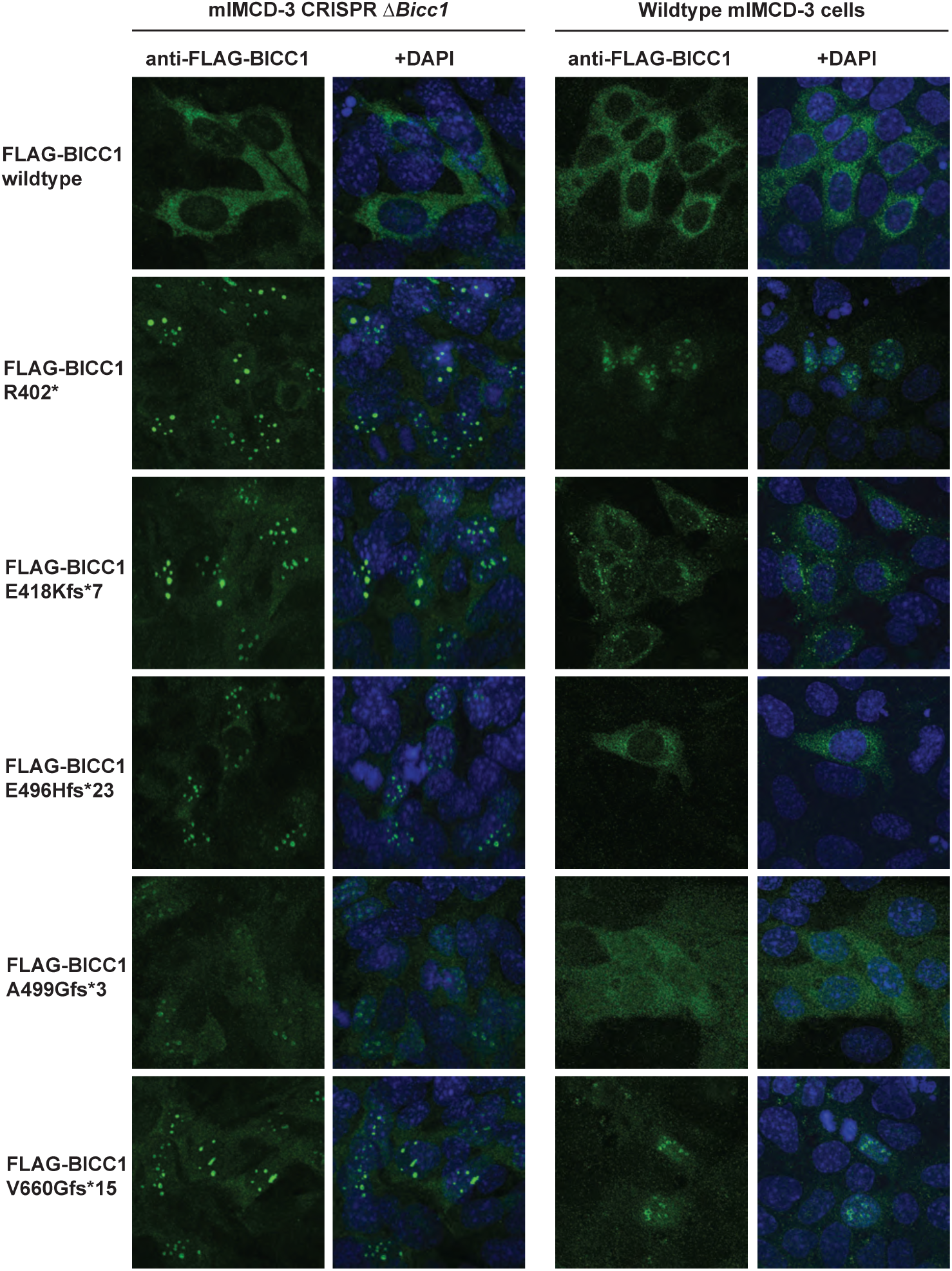
Expression of wildtype and truncated *BICC1* gene products in native and CRISPR/cas9 Δ*Bicc1* mIMCD-3 cells. **A–B–** anti-FLAG immunofluorescence of BICC1 isoforms in lentivirally transduced native (A) and CRISPR/cas9 Δ*Bicc1* mIMCD-3 cells (B). While full-length BICC1 protein distributes across the cytosol, all truncation variants demonstrate a granular localization pattern. Granular localization is more pronounced in Δ*Bicc1* cells, while expression in native mIMCD-3 cells shows an overlap between granules over a diffuse cytosolic background.

### Protein-protein interactions with SAM-domain proteins

The only available structural study suggests a model in which BICC1 molecules form multimers, mediated by stacking of their C-terminal SAM-domains^20^. The BICC1 oligomer may further attract the SAM-domain proteins, ANKS3 and ANKS6. Since our described truncation variants lack the SAM-domain, we sought to test if truncated BICC1 protein variants exhibit altered interactions with wildtype BICC1, ANKS3 and ANKS6 protein. To this end, we transiently expressed truncation-encoding *BICC1* variant cDNAs along with differentially FLAG- or Streptavidin-tagged *BICC1-, ANKS3-* and *ANKS6* wild-type cDNA in HEK293T cells, and examined protein-protein interactions through co-immunoprecipitation (co-IP) and western blot. As shown in **Figure 6A-C**, the wild-type BICC1 control protein co-precipitated with itself, as well as with ANKS3 and ANKS6. Although the BICC1 variants differ only minimally by the position of the truncation within the linker region, we found considerable variation in the binding to the tested interaction partners, including total loss or significant increase in binding affinity: The Arg402* variant exhibited wildtype-like affinity to full-length BICC1 protein, while binding to ANKS3 and ANKS6 was increased. The Glu418Lysfs*7, Ala499Glyfs*6 and Val660Glyfs*15 variants demonstrated partial or complete loss of binding to wild-type BICC1 and ANKS3, while interaction with ANKS6 remained preserved. In contrast, the Thr496Hisfs*23 truncation variant showed massively enhanced binding to all three examined binding partners, exceeding the affinity of the wild-type BICC1 protein. All protein-protein interaction observations are summarized in **Table 3**. Since binding to RNA may play a role in multimer stability, we conducted all co-IP experiments with and without inclusion of RNAse A. We found that all binary protein-protein interactions took place irrespective of the presence or absence of RNAse A, and digestion of RNA only minimally affected wild-type BICC1 oligomerization and ANKS6 binding to Thr496Hisfs*23 and Val660Glyfs*15 isoforms.

**Figure 6.**
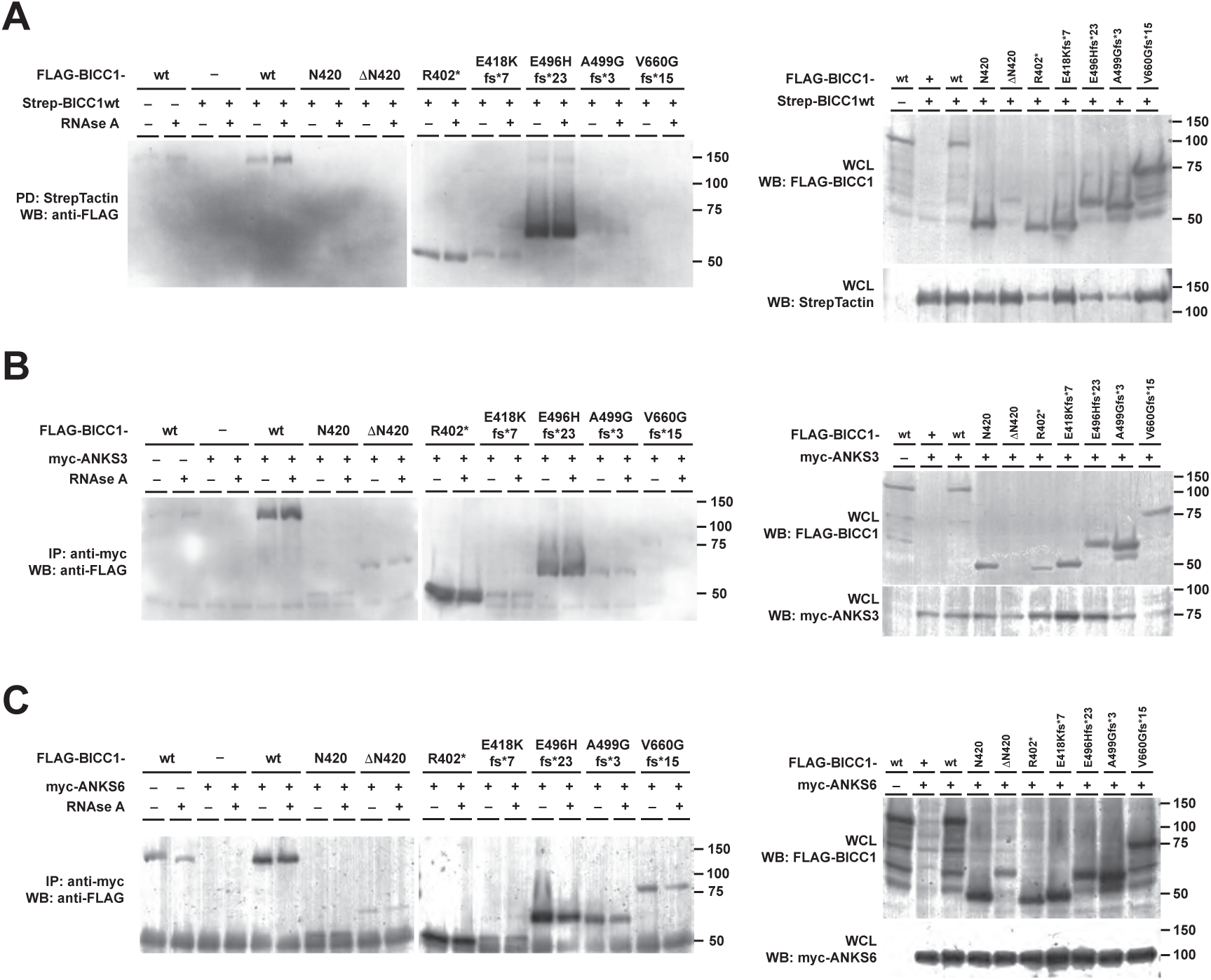
Interaction of wt BICC1 and truncation mutants with full-length BICC1, ANKS3 and ANKS6. All cDNAs were transiently transfected into HEK293T cells. We expressed differentially tagged ANKS3, ANKS6, as well as wild-type and mutant BICC1 cDNAs. Beyond the ADTKD-related variants, we also examined two engineered BICC1 truncation variants, N420 (K-homology domains) and ΔN420 (N-terminal truncation of the K-homology domains). **A–** Transient co-transfection of the indicated FLAG-tagged BICC1 variants with full-length Strep-tagged BICC1 cDNA. StrepTactin-pulldown in the presence or absence of RNAse A. The left panels show the co-precipitated BICC1 truncation mutants upon detection with anti-FLAG antibody. Despite the lack of the SAM-domain, R402*, E418Kfs*7 and E496Hfs*23 truncation products bind to the wild-type BICC1 protein. The right panel shows the Strep- and FLAG-tagged proteins in the whole cell lysates (WCL). **B–C–** Transient co-transfection of FLAG-tagged BICC1 variants with full-length myc-tagged ANKS3 (**B**) and ANKS6 (**C**) cDNA, with and without RNAse A. The left panels show the co-precipitated BICC1 variants after immunoprecipitation with anti-myc agarose and Western blot with anti-FLAG antibody. Interactions with ANKS3 and ANKS6 are detectable despite the absence of the SAM-domain in truncation variants R402*, E496Hfs*23, A499Gfs*3 and V660Gfs*15. The right panels show FLAG- and myc-tagged proteins in the whole cell lysates.

**Table 3.**
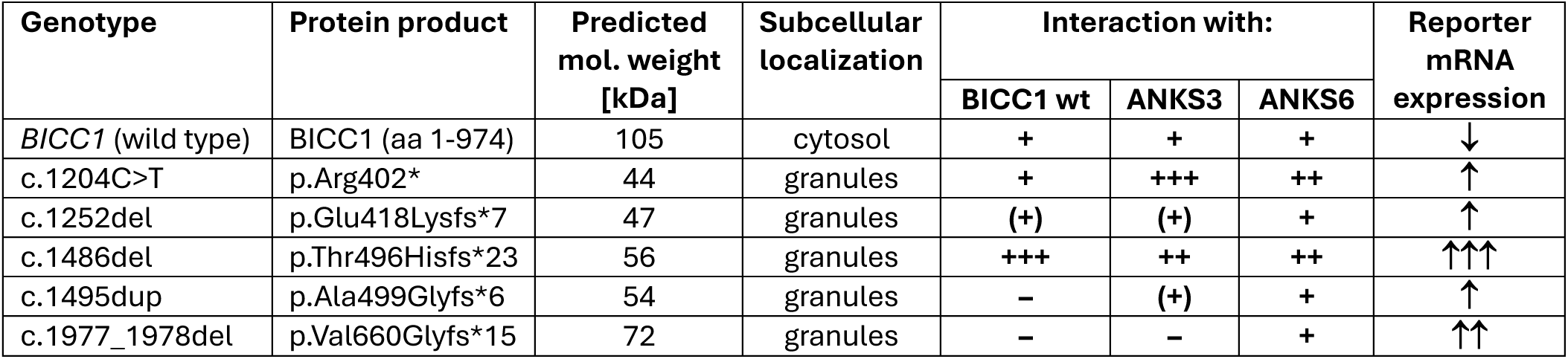
Summary of BICC1 protein isoforms and their properties.

### BICC1 truncation variants increase expression of target genes

BICC1 is a posttranscriptional regulator of specific RNA targets, acting through KH-domain-mediated binding of RNA substrates and subsequent degradation through determinants associated with the interdomain linker region^21^. Since all identified truncation variants affect the interdomain linker, we hypothesized that these truncations may affect substrate mRNA expression. Among the few known specific mRNA targets of BICC1 is *Dand5*, binding to the KH-domains through its 3’-untranslated region^25,26^ (UTR). We adapted the firefly luciferase reporter assay of Rothé et al.^27^, cloning a firefly luciferase open reading frame, followed by a stop codon and by 144 nucleotides of the *Xenopus tropicalis Dand5*-3’UTR. We transiently transfected this reporter construct into native mIMCD-3 cells, as well as into our mIMCD-3 CRISPR/cas9 *Bicc1* knock-out cell line. Consistent with its described role as a posttranscriptional repressor, *Bicc1* knockout cells exhibited a significantly increased expression of the luciferase reporter. Lentiviral add-back transduction of *Homo sapiens* wild-type *BICC1* cDNA into *Bicc1* knock-out cells resulted in signal suppression, and thus in the rescue of the knockout phenotype (**Figure 7A**). This confirms that the measured signal was indeed *BICC1*-mediated, and not an off-target effect in the CRISPR/cas9-manipulated cell line. To systematically interrogate pathogenic *BICC1* variants for their signaling activity, we transiently co-transfected the *Dand5*-3’UTR-luciferase reporter construct into *Bicc1* knockout mIMCD-3 cells, along with wild-type or mutant *BICC1*, tagged with an N-terminal *Renilla* luciferase open reading frame as a ratiometric probe. This allowed us to mimic any *BICC1* variant genetic background and analyze quantitative firefly luciferase reporter expression, normalized for the stoichiometric amount of the respective *BICC1* isoform. When expressed in *Bicc1* knock-out cells, all truncation variants except the Arg402* truncation increased the expression of the reporter construct, compared to the wild-type isoform. It is noted that this experimental setup mimicked a homozygous state in which the respective *BICC1* truncation was the only isoform present. To model a heterozygous genetic background, we co-expressed a 1:1 mix of wild-type and mutant *BICC1* cDNAs in *Bicc1* knock-out cells, along with the *Dand5*-3’UTR-luciferase reporter. Interestingly, when co-expressed with wildtype *BICC1* cDNA, all variants, including the Arg402* truncation, increased the expression of the reporter construct (**Figure 7B**). We also tested *Dand5*-3’UTR-luciferase reporter activity in HEK293T cells that expressed endogenous wild-type BICC1 gene product, and we reproduced the same findings as in the *Bicc1* knock-out mIMCD-3 cell assay modeling the heterozygous condition (**Figure 7C**). We conclude from the transcriptional reporter assay that all variants act in a dominant mode of pathogenicity, with the Arg402* truncation likely functioning as a dominant-negative variant, while the other ones likely demonstrate a gain-of-function phenotype.

**Figure 7.**
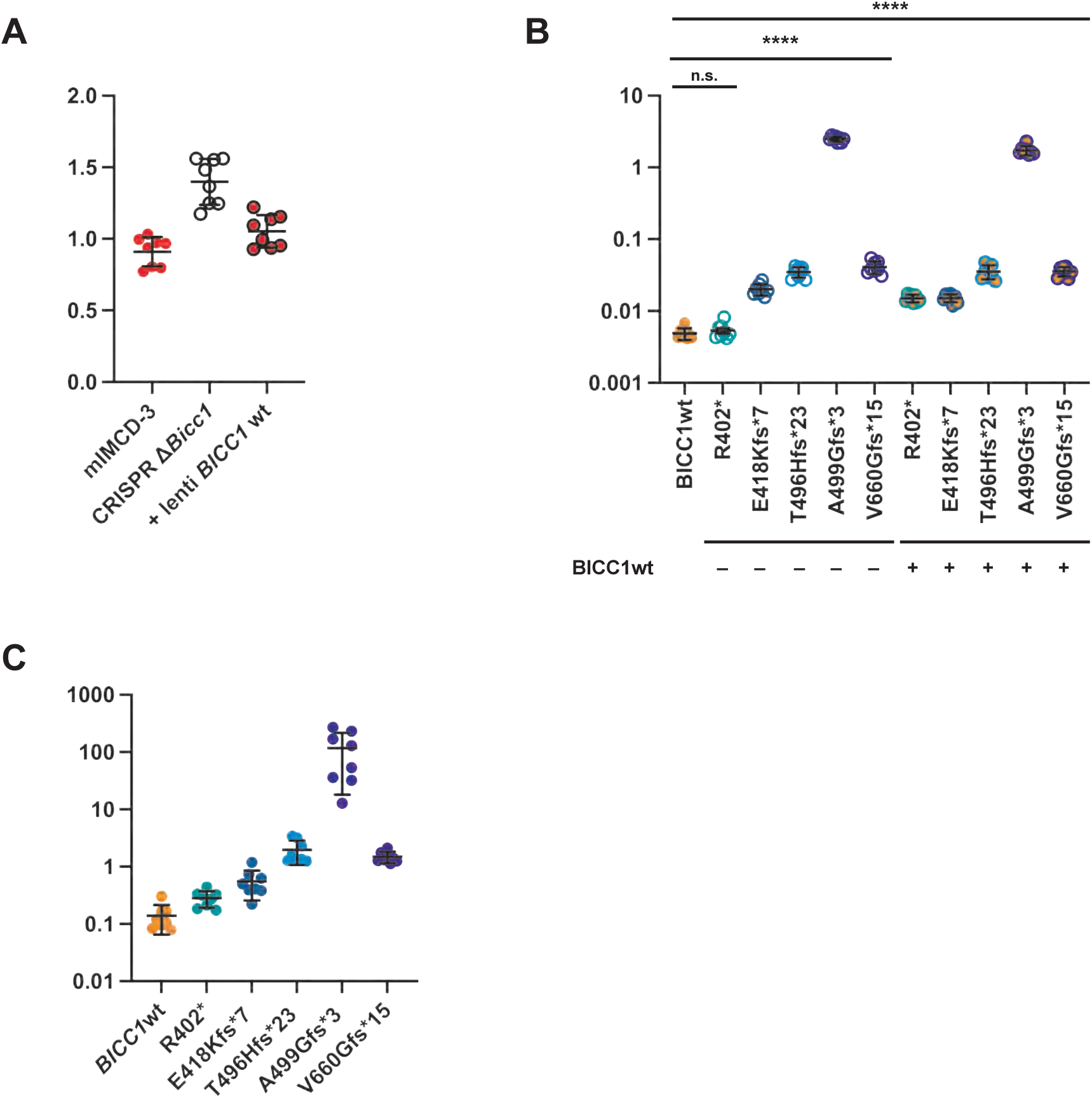
Dual-Luciferase reporter assays of BICC1 posttranscriptional regulation of *Dand5*-mRNA expression. **A–** mIMCD-3 cells exhibit a lower level of Dand5-3’UTR-mediated Luciferase activity than CRISPR/cas9 Δ*Bicc1* mIMCD-3 cells. This phenotype is rescued by lentiviral add-back expression of BICC1 full-length cDNA. **B–** Transient transfection of CRISPR/cas9 Δ*Bicc1* mIMCD-3 cells with different *BICC1* isoforms. All variants except R402X exhibit increased Luciferase activity, when compared to the full-length *BICC1* cDNA. All variants show enhanced Luciferase activity upon co-expression with full-length *BICC1* cDNA. **C–** Transient expression of BICC1 variant cDNAs in HEK293T cells recapitulate the same reporter activity as in mIMCD-3 cells.

## Discussion

In this study, we present the discovery of heterozygous truncating *BICC1* variants as a novel cause of ADTKD. ADTKD-*BICC1* is characterized by slowly progressive CKD leading to KF in the 6^th^-7^th^ decade of life, and by the lack of distinctive histopathologic features beyond tubular atrophy, interstitial fibrosis and secondary focal glomerulosclerosis. Our claim of causality is supported by cumulative evidence from segregation analysis, protein-protein interaction studies and a ratiometric BICC1 activity assay.

The presented genetic analysis highlights the identification of eight families carrying distinct heterozygous truncating *BICC1* alleles, with one recurrent variant (c.1204C>T) arising independently in three unrelated kindreds. All identified variants showed complete segregation with the CKD phenotype in 15/17 genetically tested individuals, while two individuals of young age have likely not developed clinically relevant disease, yet. In 20 additional first-degree relatives who were not amenable for genetic testing, the segregation of their CKD/KF-phenotype was consistent with an autosomal-dominant inheritance pattern.

We next sought to determine how truncating variants in *BICC1* may cause dominant disease, basing our mechanistic study on the known structure-function model of BICC1. The known crystal structure demonstrates organization of BICC1 protein into large oligomers through SAM-/SAM-domain stacking, and recruitment of the SAM-domain proteins, ANKS3 and ANKS6^20^. These oligomers are believed to spatially organize binding, sequestration and degradation of specific target RNAs through BICC1 K-homology- and interdomains^27^. We initially hypothesized that truncating mutations would result in expression of functionally insufficient BICC1 isoforms that, due to the lack of the SAM-domain, fail to incorporate into oligomers with the wild-type protein, but compete for binding to the same RNA substrates. Consequently, truncated BICC1 variants would act in a dominant-negative manner.

Intriguingly, the examined BICC1 truncation products exhibited variable protein-protein interaction properties: While three species (E418Kfs*7, A499Gfs*3 and V660Gfs*15) showed reduced or absent binding, two other truncations (R402* and E496Hfs*23) appeared to bind to full-length BICC1, ANKS3 and ANKS6 even stronger than the wild type. All truncations are located within 250 amino acids of an unstructured linker region that connects the densely packed KH- and SAM-domains; between some of the variants, the site of truncation differs only by a few amino acids. The high variability in protein complex formation patterns between variants is astonishing, and the model of BICC1 oligomer formation through SAM-domain stacking does not sufficiently explain our observations^20^. The incorporation of R402* and E496Hfs*23 variants into BICC1/ANKS3/ANKS6 oligomers is also consistent with a dominant-negative mode of action, since the presence of mutant protein likely interferes with the function of the native protein complex. In the reporter assay, wild-type *BICC1* repressed the translation of *Dand5*-3’UTR-tagged luciferase, whereas expression of *BICC1* truncation variants resulted in upregulation of the reporter. In conclusion, some variants exert their pathogenicity through incorporation into wild-type BICC1 oligomers, while other variants do not bind to wild-type BICC1/ANKS3/ANKS6 protein complexes.

Although this study is the first study systematically describing ADTKD caused by *BICC1* mutation, *BICC1* has already been reported in the context of human and other vertebrate kidney disease. A recent genome-wide association studies identified *BICC1* as a CKD risk gene through the cumulative effects of rare pLoF variants associated with CKD, KF, hypertension and gout^28^, while in a different study, a *BICC1* variant has been associated with longitudinal CKD progression^29^. *BICC1* was known as a causative gene in two recessive mouse models of PKD^5^, and from a case report of recessive CAKUT in humans^4^. An additional case series correlating heterozygous *BICC1* variants with a presumed dominant CAKUT phenotype in two children did not comment on any phenotypes in the parents and did not prove causality^30^. The exact pathways of *BICC1*-related pathogenesis in human recessive CAKUT and in murine recessive cystic disease remain elusive. Some mechanistic insights come from the study of genetic perturbations in various animal models: Outside the kidney, *BICC1* was shown to play a key role in the determination of left-right asymmetry at the embryonic node, through left-sided flow-induced degradation of *Dand5* mRNA^25,26^. Homozygous deletion of *Bicc1* in mice was shown to cause perinatal lethality, polycystic kidneys, congenital heart defects and randomization of L-/R-asymmetry^22,24,31^.

Strikingly, this syndrome phenocopies the mouse homozygous knockout of (or loss-of-function mutation in) *Pkd2*^32,33^*, Invs*^34,35^*, Nek8*^36^, *Nphp3*^37^, *AnksC*^7,38,39^ and *Anks3*^40,41^, suggesting that these genes act along the same pathway. In humans, homozygous or compound heterozygous mutations in *INVS (NPHP2)*^42^*, NPHP3*^43^*, NEK8 (NPHP9)*^44^ and *ANKSC (NPHP1C)*^39^ are known to cause nephronophthisis, a pediatric autosomal-recessive kidney disease whose histopathologic phenotype is indistinguishable from ADTKD. How the primary-cilium-localized gene products PKD2, INVS, NPHP3, NEK8 and ANKS6 communicate with cytoplasmic BICC1 and ANKS3 protein remains unknown. Some studies suggest a direct interaction between PKD2 and BICC1 gene products^45^, while others suggest the transduction of ciliary signals through phosphorylated ANKS6^46^, which, in turn, could shuttle between ciliary inversin compartment and cytosolic BICC1 foci. We speculate that BICC1 acts primarily in the cystosol, controlling the expression of specific substrate RNAs involved in kidney development and tissue homeostasis pathways, and that it may be the target for signals transduced by the polycystins and the ciliary inversin compartment. The nature of the translational responses downstream of BICC1 may result in normal kidney development and maintenance, structural anomalies (like in CAKUT), cystogenesis (like in PKD), or in accelerated tubular atrophy and interstitial fibrosis (like in NPHP and ADTKD).

In summary, truncating variants in *BICC1* are revealed as a novel cause of ADTKD in eight unrelated families. The mechanism of disease pathogenesis appears to be fundamentally different in ADTKD-*BICC1*, as compared to other causative genes: In ADTKD-*UMOD*, -*MUC1* and -*REN*, tubular epithelial cell damage originates from the accumulation of misfolded protein along the secretory pathway, resulting in proteotoxic stress through activation of unfolded protein response signaling^46-48^. In contrast, in ADTKD-*BICC1* the mutated gene product is a cytosolic protein regulating genes that control kidney tissue development and -homeostasis. Our discovery will hopefully lead to a better recognition and diagnosis of ADTKD. The pathogenic mechanism of ADTKD-*BICC1* is distinct from the other ADTKD subtypes, where proteotoxic stress leads to direct tubular cell injury. In its role as a key regulator of gene expression in kidney tissue, there is a unique opportunity in the study of *BICC1* as a therapeutic target, not only in ADTKD-*BICC1*, but more, in interstitial fibrosis and tubular atrophy in CKD of any etiology.

## Supporting information

Combined supplementary data

## Data Availability

All data produced in the present study are available upon reasonable request to the authors

## Acknowledgements

SK was supported by the Ministry of Education, Youth and Sports of the Czech Republic through the following projects: the MULTIOMICS_CZ (Programme Johannes Amos Comenius CZ.02.01.01/00/23_020/0008540) and the National Institute for Treatment of Metabolic and Cardiovascular Diseases (CarDia; LX22NPO5104) – both Co-funded by the European Union; the INTER-EXCELLENCE II grant LUAUS24087; and by the National Center for Medical Genomics (LM2023067), which kindly provided sequencing and bioinformatic analysis.

MŽ was supported by grant NW26-07-00090 from the Agency for Health Research of the Czech Republic. KS was supported by the project GAUK 38226 from The Charles University Grant Agency. Institutional support was provided through programs of Charles University in Prague (UNCE/24/MED/022 and Cooperatio).

AJB was supported by The Carlos Slim Health Foundation, the Black Brogan Foundation, the Rassmuss Foundation, Critical Path Institute US Food and Drug Administration Contract 75F40124C00106, CKD Biomarkers Consortium Pilot and Feasibility Studies Program funded by NIH-NIDDK (U01 DK103225) and Soli Deo Gloria.

GB was supported through NIH grants K08CA245100

PGC was supported through NIH grants K08DK123400, 1U24DK126110, and Brain Aneurysm Foundation grant BAF2023-6508450938. We thank Vanessa Costa, Irene Castrosin, John C. Tigges (BIDMC FACS core facility), Xu Xu (BIDMC confocal microscopy core facility), Lauren Francey, Calum Tattersfield and Meghan Long (Friedman and Pollak Labs at BIDMC) for their help and technical assistance. We thank J.H., D.U., L.R. and S.A. for their philanthropy.

The authors thank all participating individuals and their families. We would also like to thank Victoria Robins, former clinical research nurse at Wake Forest, for her invaluable work with families.

