## Supplementary material for "Heterozygous truncating variants in *BICC1* are a novel cause of autosomal-dominant tubulointerstitial kidney disease": Combined supplementary data

#### I. Supplementary Materials and Methods

##### Origin of plasmids and cDNA clones

pCMV-IRES-Renilla Luciferase-IRES-Gateway-Firefly Luciferase (pIRIGF) was a gift from William Kaelin (Addgene plasmid #101139; http://n2t.net/addgene:101139; RRID: Addgene_101139)[50].

pSpCas9(BB)-2A-GFP (PX458) was a gift from Feng Zhang (Addgene plasmid #48138; http://n2t.net/addgene:48138; RRID: Addgene_48138)[51].

psPAX2 was a gift from Didier Trono (Addgene plasmid #12260; http://n2t.net/addgene:12260; RRID: Addgene_12260).

pMD2.G was a gift from Didier Trono (Addgene plasmid #12259; http://n2t.net/addgene:12259; RRID: Addgene_12259).

Human *BICC1* cDNA was obtained from the Dana Farber Cancer Institute ORFeome collection (#100062697).

*Xenopus tropicalis Dand5* 3'UTR was amplified by PCR from Xenopus Gene Collection XGC clone #7809275 (Horizon Discovery).

*Mus musculus Anks3* and *Rattus norvegicus Anks6* wild-type cDNA constructs were kindly shared by Dr. David Beier[7].

pCDH-UbC-MCS-EF1A-Hygro was obtained from System Biosciences (SBI #CD615B-1).

pcDNA3.1(+) was obtained from Invitrogen.

##### PCR primer sequences

| Primer ID | Sequence |
|---|---|
| sgBicc1_1-fwd | 5'-CACCGCGAGCGCAGCACCGACTCGC-3' |
| sgBicc1_1-rev | 5'-AAACGCGAGTCGGTGCTGCGCTCGC-3' |
| MmBicc1_Exon1-fwd | 5'-CGCGCCGAGCCACATC-3' |
| MmBicc1_Exon1-rev | 5'-CCTGTGGCGGACTTGGGAGAGG-3' |
| MmBicc1_Exon1nested-fwd | 5'-TCGCACCATGGCCTCGCAG-3' |
| MmBicc1_Exon1nested-rev | 5'-TAGGAGCAGGGGCATGCCTAC-3' |
| hsBICC1-BamHI-fwd | 5'-TCAGGATCCATGGCCGCCCAGGGAG-3' |
| hsBICC1-NotI-rev | 5'-CACTGCGGCCGCTACCAGCGGCCACTGACAC-3' |
| hsBICC1-425-BamHI-fwd | 5'-TCAGGATCCATGGCAACCAGTCCATCCCCAGC-3' |
| hsBICC1-N420-NotI-rev | 5'-CACTGCGGCCGCTAACTGCTTTCAAGTCCGAGGAGAC-3' |
| hsBICC1-R402*-NotI-rev | 5'-GACTGCGGCCGCTACTCAACACTTTTCACAATCACAGACT-3' |
| hsBICC1-1252del-NotI-rev | 5'-GTCTGCGGCCGCTATGGTAACCCCACTGCTTTAAGTCCGAGG-3' |
| hsBICC1-1486del-fus-fwd | 5'-TCTGGTACACCCAGCCCCCATTATGGGCAC-3' |
| hsBICC1-1486del-fus-rev | 5'-AGTGGGGGTGCCCATAATGGGGGCTGGGTG-3' |
| hsBICC1-1495dup-NotI-rev | 5'-CACTGCGGCCGCTAGCAAGTGGGGGTGCCCCATAATGTGGGGCTG-3' |

| | |
|---|---|
| hsBICC1-1977_78del-fus-fwd | 5’-GTTTCCTGTGCCAAAAGGCAGACTGGAACTATTGC-3’ |
| hsBICC1-1977_78del-fus-rev | 5’-CAAAAGGCAGACTGGAACTATTGCAAGGCACGAAA-3’ |
| hsBICC1-T496Hfs*23-NotI-rev | 5’-GTCTGCGGCCGCTATAAGGTGTGGTATAGCAGAAAAACC-3’ |
| hsBICC1-V660Gfs*15-NotI-rev | 5’-GACTGCGGCCGCTAGTGCTGTGTAAGTGTGAGTTTTTC-3’ |
| Renilla-Luc-NheI-fwd | 5’-TCAGCTAGCATGACTTCGAAAGTTTATGATCCAGAAC-3’ |
| Renilla-Luc-dStop-BamHI-rev | 5’-ACTGGATCCTTGTTCATTTTTGAGAACTCGCTC-3’ |
| Firefly-Luc-NheI-fwd | 5’-TCAGCTAGCATGGAAGACGCCAAAAACATAAAG-3’ |
| Firefly-Luc-dStop-BamHI-rev | 5’-ACTGGATCCCACGGCGATCTTTCCGCCC-3’ |
| XtDand5-3UTR-BamHI-fwd | 5’-TCAGGATCCCGCCCCAACCCCACTAAATG-3’ |
| XtDand5-3UTR-KpnI-rev | 5’-ACTGGTACCTTAAATAAAGTCGTCAAGTCGTTGGC-3’ |
| MmAnks3-BamHI-fwd | 5’-CGGGATCCATGTCTGAGCTCAGCGATG-3’ |
| MmAnks3-NotI-rev | 5’-ATAGTTTAGCGGCCGCTACGGTTCCCGCCATTTC-3’ |
| RnANKS6-BamHI-fwd | 5’-CGGGATCCATGGGCGAGGGCGCG-3’ |
| RnANKS6-NotI-rev | 5’-ATAGTTTAGCGGCCGCTACCTCCTGCTCGACACTGTTTC-3’ |

#### Cell lines, culture media, reagents and supplements

| **Cell lines** | | |
|---|---|---|
| **Cell line** | **origin** | **ID** |
| HEK293T | ATCC | CRL-1573 |
| mIMCD-3 | ATCC | CRL-2123 |
| mIMCD-3 Δ*Bicc1* #B1B3 | this study | *see genotyping results below* |
| **Cell culture media and reagents** | | |
| **Reagent** | **Manufacturer** | **ID** |
| DMEM | Corning | #10-013-CV |
| DMEM/F-12 1:1 | Corning | #10-090-CV |
| Trypsin/EDTA | Corning | #25-053-CI |
| FBS | Corning | #35-010-CV |
| Penicillin/Streptomycin | Corning | #30-002-CI |
| OptiMEM I Reduced Serum Medium | Gibco | #51985034 |
| **Reagent kits and hardware** | | |
| **Item** | **Manufacturer** | **ID** |
| DNeasy Blood and tissue kit | QIAGEN | #69504 |
| Dual-glo® assay kit | Promega | #E2920 |
| Western Lightning® Plus ECL | Perkin Elmer | #NEL105001EA |
| Protein Assay Dye concentrate | Bio-Rad | #5000006 |
| Pierce Micro-spin columns | Thermo Scientific | #89879 |
| Amersham Protran 0.2 μm Nitrocellulose membrane | Cytiva | #10600001 |
| Vectashield | Vector Labs | #H-1000 |
| **Enzymes for molecular cloning** | | |
| **Enzyme** | **Manufacturer** | **ID** |
| BamHI-HF | New England BioLabs | R3136S |
| KpnI-HF | New England BioLabs | R3142S |

| | | |
|---|---|---|
| NheI-HF | New England BioLabs | R3131S |
| NotI-HF | New England BioLabs | R3189S |
| T4-DNA-Ligase | New England BioLabs | M0202S |
| Phusion High Fidelity DNA polymerase | New England BioLabs | M0530S |
| **Other biochemicals and reagents** | | |
| Bovine serum albumin (BSA) | Sigma-Aldrich | A7906 |
| Biotin | Sigma-Aldrich | B4639 |
| DAPI | Sigma-Aldrich | D9542 |
| Ethanolamine | Sigma-Aldrich | E9508 |
| 3x FLAG peptide | Sigma-Aldrich | F4799 |
| Formaldehyde | Fisher Scientific | F79500 |
| 3x myc peptide | Sigma-Aldrich | M2435 |
| Hexadimethrine bromide | Sigma-Aldrich | H9268 |
| Puromycin | Thermo Scientific | #227420100 |
| StrepTactin-HRP | IBA Life Sciences | #2-1502-001 |
| Triton X-100 | Thermo Scientific | #28314 |

| **Antibodies and affinity media** | | | |
|---|---|---|---|
| **Antibody** | **Manufacturer** | **ID** | **Titer** |
| Anti-FLAG-M2 agarose | Sigma Aldrich | A2220 | n/a |
| Anti-myc-agarose | Sigma Aldrich | A7470 | n/a |
| StrepTactin®XT-4Flow agarose | IBA Life Sciences | #2-5010-010 | n/a |
| Anti-BICC1 | LSBio | LS-C344204 | 1:50 (IHC) |
| Anti-FLAG M2 | Sigma Aldrich | F1804 | 1:5000 (WB); 1:4000 (IF) |
| Anti-FLAG (rabbit) | Sigma Aldrich | F7425 | 1:5000 (WB) |
| Anti-myc 9E10 | Sigma Aldrich | M5546 | 1:5000 (WB) |
| Anti-myc (rabbit) | Sigma Aldrich | C3956 | 1:5000 (WB) |
| Anti-Nucleolin | Sigma Aldrich | N2662 | 1:4000 (IF) |
| Anti-Edc4 (F-1) | Santa Cruz | sc-374211 | 1:200 (IF) |
| Anti-GW182 (4B6) | Santa Cruz | sc-56314 | 1:500 (IF) |
| Goat-anti-rabbit-AlexaFluor-488 | Thermo Scientific | | |
| Goat-anti-rabbit-AlexaFluor-647 | Thermo Scientific | | |
| Goat-anti-mouse-IgG1-AlexaFluor-488 | Thermo Scientific | | |
| Goat-anti-mouse-IgG1-AlexaFluor-647 | Thermo Scientific | | |

**Genotyping of mIMCD-3 Δ*Bicc1* #B1B3 cell line**

Nested PCR of the ROI in mouse *Bicc1* Exon1 and next-generation sequencing (Amplicon-EZ, Azenta) reveals three alleles, nt.79ins[T] -> p.Ser27Phefs*41, nt.80del -> p.Ser27Cysfs*68, nt.59_78del -> p.Asn20Ilefs*41, all predicted to result in early truncations. No wild-type DNA was detected.

**Long-range PCR of the Chr10:60 262 443-317 165 genomic locus in carriers of the *BICC1* c.1204C>T, p.Arg402* variant**

A 15-kb genomic DNA fragment covering the region of the *BICC1* c.1204C>T, p.Arg402* variant (Chr10:60 262 443-317 165) was amplified by LA Taq® DNA Polymerase (TaKaRa) according to the manufacturer's protocol. Amplicons were purified using magnetic beads SPRI (Beckman Coulter, Inc., Brea, CA, United States) according to the manufacturer's protocol. Purified PCR products were quantified using Qubit 2.0 Fluorometric Quantitation (Beckman Coulter, Inc.) with the Qubit™ 1x dsDNA HS Assay Kit (Thermo Scientific).

Samples were sequenced using the MinION Oxford NANOPORE platform (Oxford Science Park, United Kingdom) according to the manufacturer's protocol and Ligation sequencing amplicons—Native Barcoding Kit 24 V14 and Flongle Flow Cell (R10.4.1).

FASTQ files were aligned to the human genome reference sequence (hg19) using minimap2 (v. 2.24) in splice mode, subsequently converted to .bam format and sorted using Samtools (v1.15.1). Reads were visualized in IGV.

#### II. Supplementary Figure Legends

**Supplementary Figure S1 | Independent occurrence of the p.Arg402* truncating variant in the *BICC1* in three unrelated families with chronic kidney disease**

**A–** Schematic representation of the analyzed ~15 kb *BICC1* locus on chromosome 10. Genomic coordinates indicate the positions of used primers (Hg19; Chr10:60,548,141–60,562,912). **B–** IGV diagrams showing SNPs identified within the analyzed *BICC1* region. Distinct combination of SNPs (haplotypes) observed in three analyzed families F3 (individuals II.1 and II.4), F6 (individual_pt) and F8 (individual III.8) demonstrate the independent origin of the truncating variant p.Arg402* (c.1204C>T) in three unrelated kindreds. **C–** Identified SNPs, including genomic position, reference and alternate alleles, dbSNP identifiers, transcript annotation (NM_001080512.3), predicted amino acid change, exon localization, and population allele frequencies from the 1000 Genomes Project and gnomAD. The p.Arg402* truncating variant is highlighted in red.

**Supplementary Table 1 | Complete list of clinical characteristics in all documented individuals from Families 1-8**

### Supplementary Figure S1

**A**

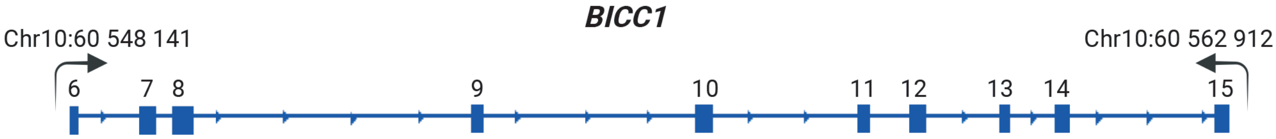

**B**

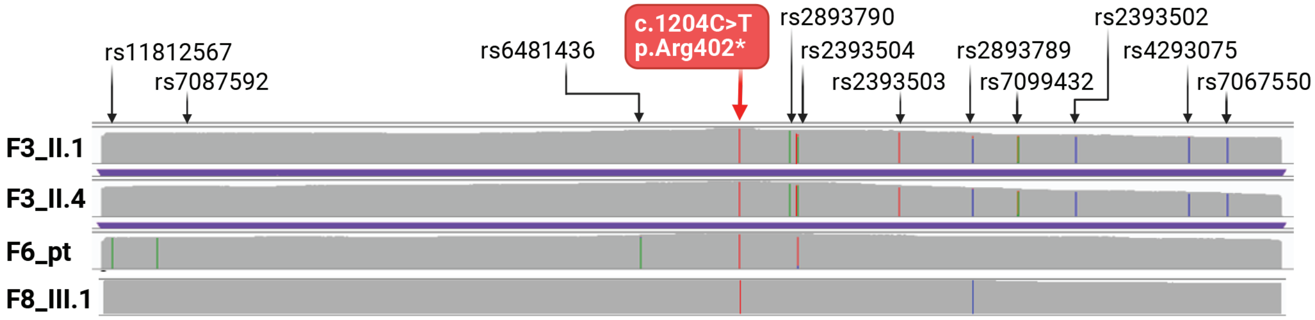

**C**

| Gene | Position | Ref | Alt | dbSnp | Transcript | AA Change | Nucleotide | Exon | Region | 1000 genomes | gnomAD (Max) |
|---|---|---|---|---|---|---|---|---|---|---|---|
| BICC1 | 60548276 | G | A | rs11812567 | NM_001080512.3 | N/A | c.600+93G>A | 6 | Intronic | 53.035% | 57.639% |
| BICC1 | 60548833 | G | A | rs7087592 | NM_001080512.3 | N/A | c.+600+650G>A | 6 | Intronic | 47.379% | 50.569% |
| BICC1 | 60554892 | C | T | rs6481436 | NM_001080512.3 | N/A | c.1180-1208C>T | 10 | Intronic | N/A | 0.02% |
| BICC1 | 60556124 | C | T | N/A | NM_001080512.3 | p.Arg402* | c.1204C>T | 10 | Exonic | N/A | N/A |
| BICC1 | 60556751 | G | A | rs2893790 | NM_001080512.3 | N/A | c.1366+465G>A | 10 | Intronic | 38.678% | 56.944% |
| BICC1 | 60556859 | G | A | rs2393504 | NM_001080512.3 | N/A | c.1366+573G>A | 10 | Intronic | 59.824% | 65.509% |
| BICC1 | 60558125 | A | T | rs2393503 | NM_001080512.3 | N/A | c.1367-34A>T | 11 | Intronic | 61.541% | 77.638% |
| BICC1 | 60559047 | G | C | rs2893789 | NM_001080512.3 | N/A | c.1725+35G>C | 12 | Intronic | 79.692% | 87.309% |
| BICC1 | 60559615 | G | A | rs7099432 | NM_001080512.3 | N/A | c.1726-339G>A | 13 | Intronic | 62.839% | 76.036% |
| BICC1 | 60560334 | G | C | rs2393502 | NM_001080512.3 | N/A | c.1858+248G>C | 13 | Intronic | 61.341% | 75.806% |
| BICC1 | 60561751 | T | C | rs4293075 | NM_001080512.3 | N/A | c.2015+945T>C | 14 | Intronic | 74.920% | 81.447% |
| BICC1 | 60562236 | T | C | rs7067550 | NM_001080512.3 | N/A | c.2016-601T>C | 15 | Intronic | 65.435% | 76.851% |

**Supplementary Table 1 | Documented clinical information of individuals from families with *BICC1* pLOF variants**

| Family | Pedigree | Genetic Diagnosis | Clinical Diagnosis | Age at eGFR or at KF | eGFR [ml $min^{-1}$] | Sex | Ultrasound | Additional Clinical Characteristics |
|---|---|---|---|---|---|---|---|---|
| 1 | I.1 | Not tested | Affected | 70s | KF | F | | None |
| | II.1 | Not tested | Affected | 60s | KF | F | | None |
| | II.4 | Not tested | Unknown | Unknown | Unknown | U | | Pancreatic cancer |
| | III.1 | p.Val660Glyfs*15 | Affected | 60s | KF | M | No | Proteinuria (self report), enuresis, hematologic cancer, Parkinson's disease, cataracts |
| | III.2 | p.Val660Glyfs*15 | Affected | 60s | 49 | M | RK:11cm; LK:12cm; No cysts | Proteinuria (ACR> 100), hypertension |
| | III.4 | p.Val660Glyfs*15 | Asymptomatic | 60s | 65 | F | No | T2DM, coronary and cerebrovascular disease, hypertension, cataracts |
| | III.5 | p.Val660Glyfs*15 | Affected | 60s | 27 | M | No | T2DM, Proteinuria (mild), hematuria, hypertension |
| | III.7 | Wild type | Healthy | 50s | 62 | F | No | None |
| | III.8 | Wild type | Healthy | 60s | 103 | M | No | None |
| | III.9 | Wild type | Healthy | 60s | 80 | F | No | None |
| | IV.2 | Not tested | Unknown | Unknown | Unknown | U | | Renal hypoplasia, urinary reflux requiring reimplantation during infancy |
| | IV.3 | p.Val660Glyfs*15 | Asymptomatic | 20s | 101 | F | No | None |
| | IV.4 | Wild type | Asymptomatic | Unknown | Unknown | F | No | None |
| | IV.5 | Wild type | Asymptomatic | Unknown | Unknown | M | No | None |
| | IV.6 | p.Val660Glyfs*15 | Asymptomatic | 30s | 122 | F | No | None |
| 2 | I.1 | Not tested | Affected | 80s | KF | F | | None |
| | II.1 | Not tested | Affected | 80s | KF | F | | T2DM, thyroid cancer, severe hearing loss, hypertension, glaucoma, COPD, hypertrophic cardiomyopathy, MI |
| | II.2 | Wild type | Healthy | 80s | 61 | M | No | None |
| | II.3 | Wild type | Healthy | 80s | 84 | F | No | None |
| | II.4 | Wild type | Healthy | 90s | 60 | F | No | None |
| | II.6 | Not tested | Affected | 80s | KF | M | | None |
| | III.1 | p.Ala499Glyfs*6 | Asymptomatic | 50s | 67 | M | No | |
| | III.2 | p.Ala499Glyfs*6 | Affected | 50s | KF | M | RK:11.1cm; LK:11cm; No cysts; Echogenic | Hypertension |

| | | | | | | | | |
|---|---|---|---|---|---|---|---|---|
| 3 | I.1 | Not tested | Affected | 70s | KF | F | | None |
| | II.1 | Not tested | Affected | 70s | KF | F | | None |
| | II.3 | Not tested | Unknown | Unknown | Unknown | U | | Death in 80s |
| | II.5 | Not tested | Affected | 30s | KF | M | | Gout, heart failure |
| | III.1 | Not Tested | Unknown | Unknown | Unknown | U | | Died from substance use disorder and additional comorbidities undisclosed. |
| | III.3 | Not tested | Affected | 60s | KF | M | | None |
| | III.4 | Not tested | Affected | 70s | KF | M | | None |
| | III.8 | p.Arg402* | Affected | 70s | KF | M | No | Enuresis |
| | III.10 | Not tested | Affected | 60s | KF | F | -- | None |
| | III.11 | p.Arg402* | Affected | 70s | 25 | F | No | Gout, followed by a hematologist |
| | IV.1 | p.Arg402* | Affected | 70s | 9 | F | No | Elevated SUA. hypertension |
| | IV.3 | Not tested | Unknown | Unknown | Unknown | U | | Diabetes of unknown type in childhood, death from ketoacidosis in 30s |
| | IV.4 | Not tested | Affected | 50s | 57 | F | | Kidney infections since childhood, surgery for kidney infections in childhood, chronic UTIs as an adult. |
| | IV.5 | Not tested | Affected | Age not given | KF | F | | Diabetes of unknown type in childhood, COVID-19 infection led to ESRD. History of potential stroke. |
| | IV.7 | Wild type | Healthy | 50s | 70 | M | No | None |
| | IV.9 | Not tested | Unknown | Unknown | KF | U | | T1DM, sees a nephrologist |
| 4 | I.1 | Not tested | Affected | 80s | KF | F | | Diagnosed with renal and hepatic cysts, hyperparathyroidism, hypertension, neuropathy |
| | II.1 | Glu418Lysfs*7 | Affected | 60s | 32 | F | RK:9cm; LK:9cm; Simple renal cysts | Hepatic cysts, distal sensorimotor neuropathy, primary hyperparathyroidism |
| | II.2 | Glu418Lysfs*7 | Affected | 60s | KF | M | RK:10cm; LK:11.3cm; RK cortical cyst 2x3cm | Hyperkalemia, gout, hypertension |
| 5 | II.1 | Thr496Hisfs*23 | Asymptomatic | 40s | 87 | M | | Proteinuria (mild) |
| 6 | | p.Arg402* | Unknown | Unknown | Unknown | U | | No clinical information available |
| 7 | I.1 | Not tested | Affected | 60s | KF | M | | Horseshoe kidney |
| | II.1 | p.His516Thrfs*14 | Affected | 50s | KF | M | | Proteinuria (mild), hypertension |
| 8 | II.1 | Not tested | Unknown | Unknown | Unknown | U | | T2DM, hypertension |
| | III.1 | p.Arg402* | Affected | 50s | 41 | M | Bilateral small | Gout, hypertension, trace hematuria |

| | | | | | | | | |
|---|---|---|---|---|---|---|---|---|
| | | | | | | | kidneys, few cysts | |
| | III.2 | Not tested | Unknown | Unknown | Unknown | U | | Gout |

Abbreviations used in Supplementary Table 1. Chronic obstructive pulmonary disease (COPD), kidney failure (KF), serum creatinine (SCr), right kidney (RK), left kidney (LK), urine albumin creatinine ratio (ACR), myocardial infarction (MI), serum uric acid (SUA), Type 1 diabetes mellitus (T1DM), Type 2 diabetes mellitus (T2DM), unknown gender (U).
